# Impact of Respiratory Syncytial Virus (RSV) in adults 60 years and older in Spain

**DOI:** 10.1101/2024.09.11.24313456

**Authors:** S. Jimeno, A. Peláez, Á. Calle, M. Villarreal, S. Natalini

## Abstract

Respiratory illnesses frequently lead to hospitalisation in adults aged 60 and older, especially due to respiratory virus infectious (RVIs). This study investigates hospitalisation patterns and characteristics of RVIs at HM Hospitals from October 2023 to March 2024.

We retrospectively explored hospitalisations of patients aged 60 years and older with RVIs, gathering data on demographics, clinical profiles, comorbidities, and treatments. Outcomes included hospitalisation, ICU admissions, and mortality, independent factors associated with outcomes were identified using a multi-state model.

From October 2023 to March 2024, from a total of 3,258 hospitalisation, 1,933 (59.3%) were identified as positive for RVIs. Overall, SARS-CoV-2 was the most prevalent (52.6%), followed by influenza (32.7%) and RSV (11.8%). Most RVIs involved single infections (88.2%). Hospitalisation rates increased with age for SARS-CoV-2, influenza, and RSV, with SARS-CoV-2 showing the highest rate, followed by influenza and RSV.

In the multi-state model, RSV infection significantly increased ICU admission risk (HR: 2.1, 95%, *p* = 0.037). Age on admission (HR: 1.1, 95%, p < 0.001) and Charlson score (HR: 1.4, 95%, *p* = 0.001) were associated with transitioning from admission to death. ICU to death risks included age at admission (HR: 1.7, 95%, *p* < 0.001).

RVIs in adults 60 years and older are associated with high hospitalisation and mortality rates, primarily driven by influenza and SARS-CoV-2, followed by RSV. Age and comorbidities significantly impact disease severity, emphasising the need for targeted prevention and management strategies for RSV in this vulnerable population.

**Funding:** This studio received no funding.

## Introduction

The year 2020 witnessed an unprecedented global health crisis due to the COVID-19 pandemic. In Spain, there has been a 68.5% increase in deaths related to respiratory diseases compared to 2019 (1). This increase not only led to an overall decline in life expectancy, but also raised concerns about the possible long-term persistence of this trend (2–4). However, the impact of respiratory infections extends beyond the context of the COVID-19 pandemic. Reports such as that of Heppe-Montero et al. (5), suggest that respiratory infections, including that caused by respiratory syncytial virus (RSV), among others, play an important role in morbidity and mortality, especially among elderly patients. As the world’s population ages, the morbidity and mortality associated with respiratory infections continues to increase (6).

For many years, the contribution of different viral agents to acute respiratory illnesses in elderly patients was primarily limited to studies on influenza virus. The scarcity of studies was largely due to diagnostic challenges and symptom overlap among different respiratory viruses (7). However, modern laboratory tools now allow for the identification of respiratory infectious viruses (RIVs). Since 2016, there has been increasing interest in RSV, evidenced by efforts by the World Health Organization (WHO) to strengthen global RSV surveillance systems. Nevertheless, much of the available data on RSV disease in adults over 60 years old comes from a limited number of geographic locations, mainly from the United States. Different cultures, climates, and multigenerational household dynamics may significantly influence infection rates (8, 9). There is an urgent need for broader data on infections caused by this virus in the population. In Spain, after the COVID-19 pandemic, the Acute Respiratory Infection Surveillance System (SiVIRA) emerged against influenza virus, SARS-CoV-2 and RSV, which demonstrates the growing interest in studying the different aetiologies of acute respiratory infections in adults on which we can intervene by changing their evolutionary course, through treatment with antivirals or monoclonal antibodies, and prevention with vaccines (10).

RSV is a typically human pathogen, impacting specific population groups comparable to or more than seasonal influenza (5, 11, 12). It is estimated to cause 3.4 million severe cases annually requiring hospitalisation (3). In recent years, RSV has emerged as a leading cause of acute respiratory illness, posing a high risk of severe complications in elderly individuals or those with chronic diseases (5, 9, 13–15). The gradual decline in innate and adaptive immune system effectiveness associated with aging (immunosenescence) likely contributes to severe RSV illness in the elderly (16, 17). This risk is compounded by high RSV incidence rates observed in specific age groups, such as those over 79 years (40.2 cases/100,000 population) and the 65-79 age group (1.5 cases/100,000 population) (18). Additionally, RSV does not induce long-lasting immunity, potentially causing multiple reinfections throughout life, which can be life-threatening in patients with underlying conditions (10, 19–21). RSV imposes a significant burden on healthcare systems. In Spain, RSV-related hospitalisations cost an estimated €50 million annually to the National Health System (22–24). Therefore, vaccines play a crucial role, especially in elderly patients or those with comorbidities (17).

Efforts over the past 15 years have focused on developing various RSV immunoprophylaxis options, such as long-acting monoclonal antibodies for passive immunisation of newborns and infants, and vaccines for pregnant women, children, and adults over 60 years old (10). On May 3, 2023, the US Food and Drug Administration (FDA) approved GSK’s Arexvy vaccine for individuals over 60 years old, administered in a single dose. A clinical trial involving nearly 25,000 adults over 60 demonstrated 83.0% efficacy in preventing lower respiratory tract illnesses and 94% efficacy against severe illness, regardless of RSV subtype and underlying comorbidities (16). On August 21, 2023, Pfizer’s Abrysvo vaccine was approved by the FDA for protection of newborns and young infants through active immunisation of pregnant women. On May 18, 2023, it received FDA approval for individuals over 60 years old. Currently, neither vaccine is funded by the Spanish National Health System. Furthermore, new RSV vaccines are being developed by Moderna and Bavarian Nordic, and there is promising potential in using monoclonal antibodies as a therapeutic strategy (25).

Diagnosing and preventing RSV in adult patients, especially the vulnerable, are crucial for reducing morbidity, mortality, and excessive healthcare resource use (12, 26). The aim of this study is to assess the impact of RSV infections in adults over 60 years of age in different communities within the national territory, identifying factors that may be related to greater severity of the disease and developing predictive models that can improve management. and the allocation of health resources.

## Material and methods

### Study Design

We conducted a multicenter retrospective observational study using secondary anonymized data extracted from the electronic health records (EHR) of adult patients aged 60 years and older admitted between October 1, 2023 and March 31, 2024 at 18 hospitals. university students of the HM group distributed throughout the country (Madrid: Madrid, Montepríncipe, Torrelodones, Sanchinarro, Vallés, Puerta del Sur and Rivas; León: San Francisco and Regla; Barcelona: Nou Delfos and Sant Jordi; Coruña: Modelo Belen; Santiago de Compostela: Rosaleda and Esperanza; and Málaga: Dr. Gálvez, Santa Elena, Clínica del Pilar and Complejo Hospitalario Integral Privado). All patients had a diagnosis of respiratory infection due to RIVs confirmed by PCR or antigen. The study was conducted in accordance with the Declaration of Helsinki, and approved by the Ethics Committee of HM Hospitales (protocol code 24.04.2344-GHM and date of approval: 2024/05/08).” for studies involving humans. Moreover, this research is a retrospective study using anonymized data, which involves no direct patient intervention; consequently, individual consent to participate is not necessary.

### Data Extraction

We collected sociodemographic variables such as admission and discharge dates from both regular wards and intensive care units (ICU), date of birth, gender, and mortality status. Additionally, we retrieved ICD-10 codes to extract various variables, including smoking status, alcohol consumption, associated pathologies (such as asthma, diabetes, obesity, hypertension, chronic obstructive pulmonary disease (COPD), renal failure, cardiac diseases, neoplasms), use of medical procedures (high-flow nasal cannula, oxygen therapy, invasive ventilation), and symptoms present during the episode (pneumonia, upper respiratory tract infection (URTI), lower respiratory tract infection (LRTI), acute respiratory distress syndrome). The Charlson Comorbidity Index was calculated for each patient during each admission using the Comorbidity Package (27). We also collected variables on pharmacological treatments administered during hospitalisation (antibiotics, anticoagulants, antihypertensives, antivirals, diuretics). Data were extracted and anonymised from local EHRs to protect patient confidentiality and privacy, then combined into a unified harmonised dataset.

### Statistical Analysis

First, a descriptive and comparative analysis of sociodemographic variables was conducted. Continuous variables were presented as mean and standard deviation (SD). Normality of continuous variables was assessed using the Shapiro-Wilk test or Kolmogorov-Smirnov test, while homoscedasticity was tested using Levene’s test. Parametric tests (t-test or ANOVA) or non-parametric tests (Kruskal-Wallis or Wilcoxon rank-sum test) were applied based on adherence to these assumptions. Categorical variables were compared using the chi-square test or Fisher’s exact test, as appropriate. Total admission cases were used to calculate incidence rate ratios and corresponding 95% confidence intervals (CI). Finally, to identify potential predictors of severity in RVIs-related hospital admissions, a multi-state model was constructed. Variables were selected using univariate modelling for each predictive variable available for each state (ICU admission, mortality). In addition, we assessed the proportional hazards criterion and whether in the case of continuous variables they followed a non-linear or time-dependent distribution. A significance level of *p* < 0.05 was used for all analyses. For multiple comparisons, corrections (Bonferroni method) were utilized to adjust for inflated Type I error rates. Data manipulation was performed by the function contained in the *tidyverse* package (28). Rate analyses were performed using the *epiR* package (29) and proportional hazards and multistate cox regression testing were performed using the survival and *mstate* packages (30,31), respectively. Data processing and statistical analyses were performed using R (32), version 4.3.1.

## Results

### Respiratory disease hospitalizations in elder patients

From October 1, 2023, to March 31, 2024, a total of 22,201 episodes involved hospitalizations of patients aged 60 and older, across HM Hospitals centres. Among these, 3,258 episodes (14.7%) were selected where patients were diagnosed with respiratory pathology, including: URTI, LRTI, flu, or pneumonia. All patients underwent viral detection testing (positive antigen test or PCR for viruses) during their hospital stay. Out of the total respiratory pathology diagnoses, 1,933 cases tested positive for at least one RVIs, with 31.2% of these patients positive for SARS-CoV-2, 19.4% for influenza virus, 7.0% for RSV, 6.0% for rhinovirus, 1.7% for metapneumovirus, and the remaining 4.9% positive for other RVIs such as adenovirus, bocavirus, *enterovirus,* and parainfluenza (**Table 1**). Among the total admissions (1,933 in total), 1,704 (88.2%) were attributed to a single RVIs, 152 (7.9%) to dual RVIs co-infections, and 77 (4.0%) to infections involving more than two RVIs. The impact of each RVIs of interest relative to the total number of unique episodes where any RVIs was detected was measured. First of all, SARS-CoV-2 was most prevalent (1,016; 52.6%), followed by influenza virus (632; 32.7%), and RSV (229; 11.8%).

**Table 1.** Impact of RVIs on Total Admissions for Respiratory Pathology and Positive Admissions with RVIs detection.

| Items | Admissions for respiratory pathology (*n* =3,258)<sup>2</sup> | Positives admissions for any of the RVIs of interest (*n* =1,933)<sup>3</sup> |
| --- | --- | --- |
| **Including co-infections** |  |  |
| SARS-CoV-2 | 1,016 (31.2%) | 1,016 (52.6%) |
| Influenza virus | 632 (19.4%) | 632 (32.7%) |
| RSV | 229 (7.0%) | 229 (11.8%) |
| Rhinovirus | 194 (6.0%) | 194 (10.0%) |
| Other viruses<sup>1</sup> | 160 (4.9%) | 160 (8.3%) |
| Metapneumovirus | 54 (1.7%) | 54 (2.8%) |
| **Single infections** |  |  |
| SARS-CoV-2 | 936 (28.7%) | 936 (48.4%) |
| Influenza viruses | 500 (15.4%) | 500 (25.9%) |
| RSV | 132 (4.1%) | 132 (6.8%) |
| Rhinovirus | 77 (2.4%) | 77 (4.0%) |
| Other viruses<sup>1</sup> | 40 (1.2%) | 40 (2.1%) |
| Metapneumovirus | 36 (1.1%) | 36 (1.9%) |
| **Major RSV co-infections** |  |  |
| RSV + rhinovirus + other viruses | 42 (1.3%) | 42 (2.2%) |
| Influenza + RSV | 36 (1.1%) | 36 (1.9%) |
| Influenza + RSV + rhinovirus + other viruses | 36 (1.1%) | 36 (1.9%) |
| Influenza + rhinovirus | 31 (1.0%) | 31 (1.6%) |
| COVID+ RSV+ rhinovirus + other viruses | 24 (0.7%) | 24 (1.2%) |

<sup>1</sup>The classification "other viruses" includes positive episodes for adenovirus, bocavirus, enterovirus, and parainfluenza.
<sup>2</sup>Includes only episodes with ICD-10 referring to respiratory pathology (Infectious diseases of the upper (J00-J06) and lower (J20-J22) respiratory tract, flu and pneumonia (J10-J18) or having a positive test for one of the RVIs of interest.
<sup>3</sup>Some patients may have had co-infection, so the sum does not add up to the total.

The admission rates, ICU admission rates and mortality rates with presence of RIVs were calculated per 10,000 admissions of patients aged 60 and above for the season spanning October 1, 2023, to March 31, 2024 (**Figure 1; Annex Table 1**). It was observed that SARS-CoV-2 admissions were highest in all months of the analysed season, except for January 2024, where it was surpassed in all three analyses (admission, ICU admission and mortality rates). In second place, influenza virus was consistently the second most prevalent virus throughout the season. RSV ranked third for most of the season, peaking during the months of November to January.

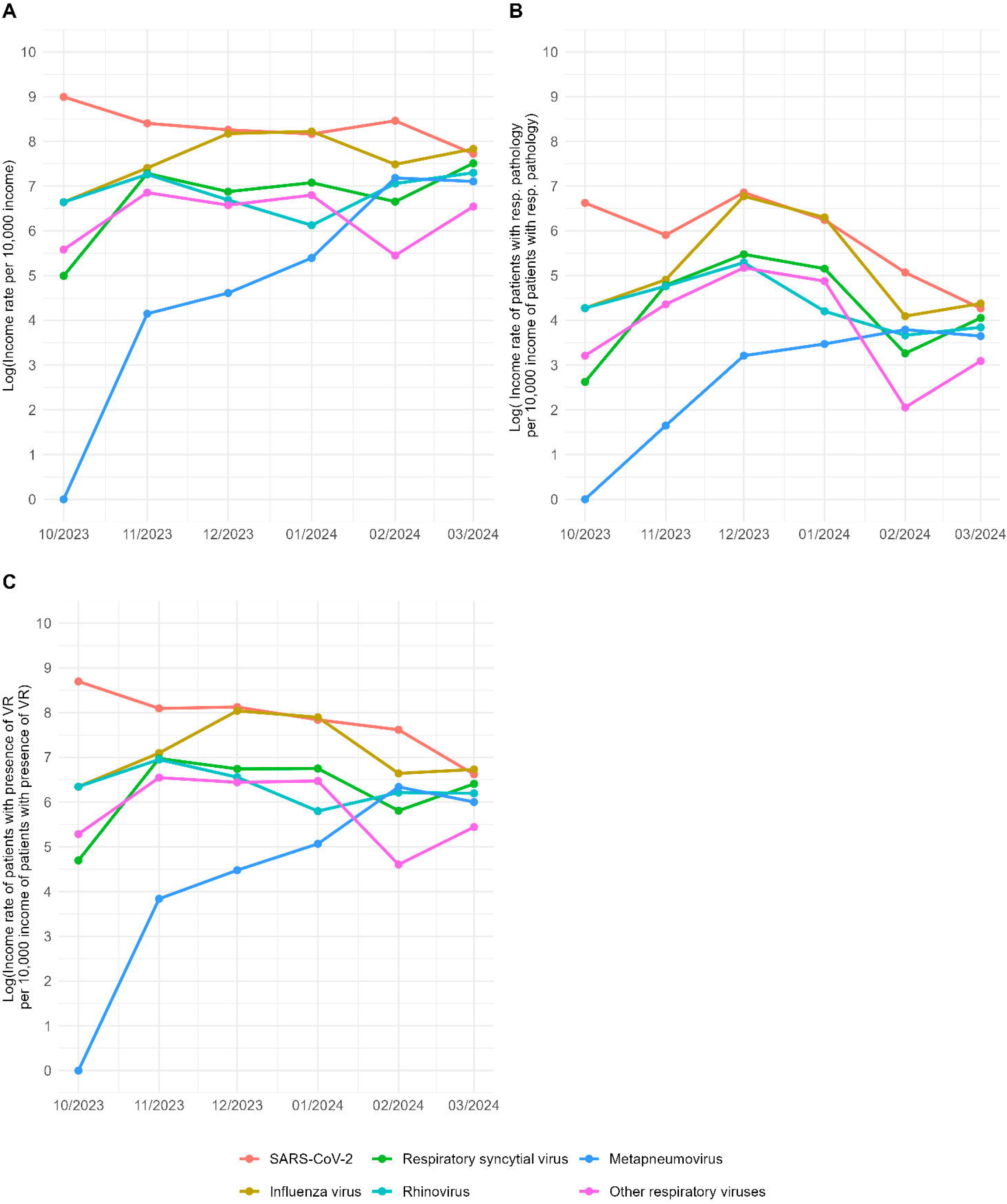

### Hospitalizations by RVIs for Each Age Group and Outcome

To assess the impact of predominant RVIs (SARS-CoV-2, influenza virus and RSV) on hospital admissions across age groups, admission rates per 10,000 admissions along with 95% confidence intervals (CI), ICU admissions, and mortality rates were analysed (**Figure 2; Annex Table 2**). The admission rate increased as the age of the group increased with any of the viral infections studied (SARS-CoV-2: 333.4 [95% CI: 295.0 – 375.2] to 651.6 [95% CI: 532.1 – 788.4]; influenza virus: 169.8 [95% CI: 142.6 – 200.7] to 518.6 [95% CI: 412.1 – 643.1] and RSV: 69.2 [95% CI: 52.2 – 90.0] to 246.0 [95% CI: 173.8 – 337.5]).

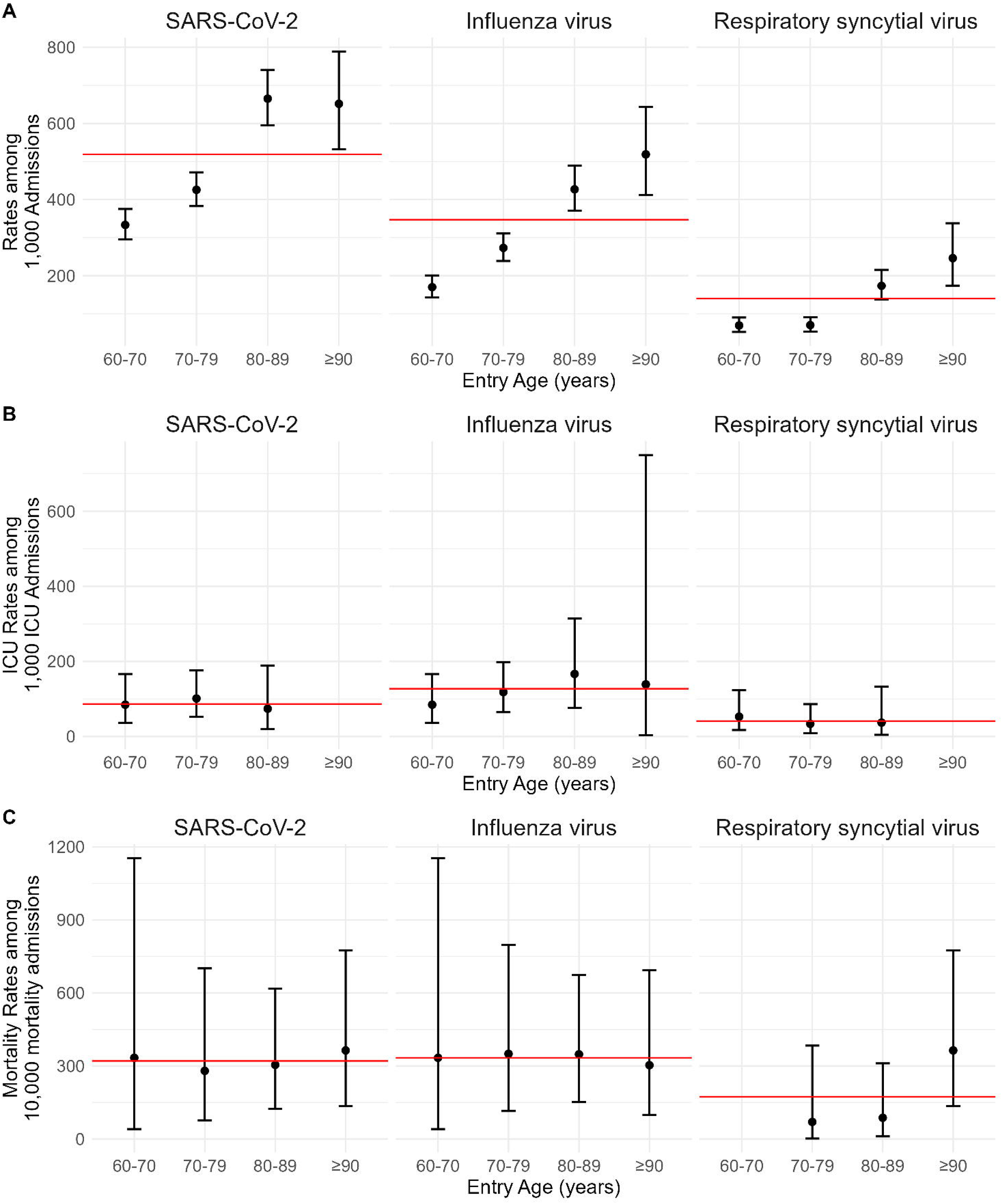

**Table 2.** Descriptive analysis of admissions with RIVs for each with single infections.

| Items | SARS-CoV-2 (n1=936) | Influenza (n2=500) | VRS (n3=132) | p-value |
| --- | --- | --- | --- | --- |
| Sex [Female]. | **522 (55.8%)↓** | **259 (51.8%)↓** | **85 (64.4%)↑** | **0.030** |
| Age at admission | **76.6 (±9.2)↓** | 77.7 (±9.5) | **79.0 (±10.4)↑** | **0.006** |
| Length of stay | **6.6 (±6.4)↓** | **8.6 (±10.4)↑** | **8.61 (±5.9)↑** | **<0.001** |
| ICU | 17 (1.8%) | 19 (3.8%) | 3 (2.3%) | 0.070 |
| ICU days | 5.2 (±7.1) | 4.7 (±8.5) | 1.3 (±0.6) | 0.435 |
| Exitus | **13 (1.4%)↓** | 11 (2.2%) | **6 (4.5%)↑** | **0.040** |
| Smoker | **64 (6.8%)↓** | **62 (12.4%)↑** | **18 (13.6%)↑** | **0.008** |
| Former smoker | 193 (20.6%) | 114 (22.8%) | 34 (25.8%) | 0.995 |
| Alcoholism | 28 (3.0%) | 26 (5.2%) | 3 (2.3%) | 0.098 |
| **Comorbidities** |  |  |  |  |
| Asthma | 41 (4.4%) | 32 (6.4%) | 7 (5.3%) | 0.428 |
| Diabetes | 157 (16.8%) | 92 (18.4%) | 22 (16.7%) | 0.468 |
| Obesity | **48 (5.1%)↓** | **64 (12.8%)↑** | **16 (12.1%)↑** | **<0.001** |
| Hypertension | 443 (47.3%) | 266 (53.2%) | 84 (63.6%) | 0.715 |
| COPD | **152 (16.2%)↓** | 117 (23.4%) | **40 (30.3%)↑** | **0.008** |
| Kidney failure | 128 (13.7%) | 84 (16.8%) | 27 (20.5%) | 0.566 |
| Heart failure | 154 (16.5%) | 95 (19.0%) | 40 (30.3%) | 0.053 |
| Neoplasia | 188 (20.1%) | 99 (19.8%) | 35 (26.5%) | 0.490 |
| Charlson Index | **2.13 (±2.9)↓** | 2.74 (±3.3) | **3.33 (±3.1)↑** | **<0.001** |
| **Use of health services** |  |  |  |  |
| High-flow nasal goggles | 3 (0.3%) | 0 (0.0%) | 0 (0.0%) | 0.199 |
| Oxygen therapy | 17 (1.8%) | 21 (4.2%) | 7 (5.3%) | 0.340 |
| VMI | 5 (0.5%) | 3 (0.6%) | 1 (0.8%) | 0.908 |
| **Symptoms** |  |  |  |  |
| Pneumonia | **81 (8.7%)↓** | **34 (6.8%)↓** | **20 (15.2%)↑** | **<0.001** |
| LRTI | **6 (0.6%)↓** | 16 (3.2%) | **8 (6.1%)↑** | **0.007** |
| URTI | **4 (0.4%)↓** | **1 (0.2%)↓** | **18 (13.6%)↑** | **<0.001** |
| Respiratory distress syndrome | 7 (0.7%) | 1 (0.2%) | 0 (0.0%) | 0.079 |
| **Treatments** |  |  |  |  |
| Antibiotics | **257 (27.5%)↓** | 240 (48.0%) | **83 (62.9%)↑** | **0.001** |
| Anticoagulants | 236 (25.2%) | 202 (40.4%) | 66 (50.0%) | 0.961 |
| Antihypertensives | 14 (1.5%) | 12 (2.4%) | 0 (0.0%) | 0.132 |
| Antivirals | **18 (1.9%)↓** | **162 (32.4%)↑** | **8 (6.1%)↓** | **<0.001** |
| Diuretics | 119 (12.7%) | 115 (23.0%) | 45 (34.1%) | 0.057 |

Data are shown as n (%), mean ( ± standard deviation). NA: Not available. LRTI: Lower respiratory tract infection, URTI: Upper respiratory tract infection. Significant differences ( $p < 0.05$ ) are indicated in bold, and the direction of the pairwise post-hoc results ( ↑ , ↓ ).

This trend significantly surpassed the average admission rate for SARS-CoV-2 (518.9), influenza virus (347.1), and RSV (139.7) in age groups 80 years and older. SARS-CoV-2 had the highest admission rate, followed by influenza virus and RSV.

Regarding ICU admission or mortality rates, there were no significant differences observed among the RVIs. However, the average ICU admission rate was higher for influenza virus (127.14) compared to SARS-CoV-2, followed by RSV (86.7 and 41.3, respectively). Specific ICU admission rates for SARS-CoV-2 infections were higher (101.4 [95% CI: 52.5 – 176.4]), and for RSV (166.7 [95% CI: 76.5 – 314.0]) in patients aged 70-79 years, while for influenza virus infections in patients aged 80-89 years (166.7 [95% CI: 76.5 – 314.0]). When evaluating mortality rates per 100,000 admissions for patients who died, the ranking by average mortality rate was as follows: influenza virus (333.5), SARS-CoV-2 (320.3), and RSV (173.5). Specific mortality rates for SARS-CoV-2 infections were higher in patients aged 70-79 years (279.7 [95% CI: 76.7 – 700.7]), for influenza virus infections in patients aged 80-89 years (347.83 [95% CI: 151.3 – 673.8]), for RSV infections in patients aged 90 years and older (363.6 [95% CI: 134.6 – 774.65]).

### Sociodemographic and Clinical Characteristics

Out of a total of 1,933 unique episodes of patients positive for RVIs, 1,721 (89.0%) were single infections. These single infections corresponded to SARS-CoV-2 (936 admissions; 54.39%), influenza virus (500; 29.1%), RSV (132; 7.7%), rhinovirus (77; 4.5%), metapneumovirus (36; 2.1%), and other minority viruses (40; 2.3%). Focusing only on the admission where main viruses were presented (1568 admissions; 936 of SARS-CoV-2, 500 of influenza virus and 132 of RSV), the sociodemographic characteristics of our sub-sample had a mean admission age of 77.1 ± 9.4 years, with a mean stay of 7.6 ± 8.2 days and admissions ending in ICU admission (39; 2.5%) had a mean stay of 4.7 ± 7.5 days.

When we focus on the characteristics of the admissions of each specific virus (**Table 2**), females were predominant across all RVIs of interest, with a significant (*p*= 0.030) higher representation in VRS admissions (85 admissions; 64.4%) and lower in SARS-CoV-2 (522;55.8%) and influenza admissions (259; 51.8%). Mean age at admission varied significantly (*p* = 0.017), being significantly lower for SARS-CoV-2 patients (76.6 ± 9.2) and higher for RSV patients (79.0 ± 10.4). Hospital length stay also showed significant differences (*p* < 0.001), with significantly longer stays for RSV (8.6 ± 5.9 days), and influenza virus (8.6 ± 10.4 days) compared to SARS-CoV-2 patients (6.6 ± 6.4 days). In terms of admission outcome, patients with RSV (6; 4.5%) had a significantly higher mortality rate than those with SARS-CoV2 (13; 1.4%). Active smokers were also significantly more prevalent in VRS (18; 13.6%), and influenza (62; 12.4%) admissions compared to SARS-CoV-2 (64; 6.8%).

Evaluation of comorbidities revealed that the proportion of admissions with obesity were significantly lower (*p*<0.001) when SARS-CoV-2 is presented (48; 5.1%) in comparison with influenza (64; 12.8%) and VRS (16; 12.1%). COPD prevalence showed significant differences (*p* = 0.008), being higher in RSV patients (40; 30.3%) and lower in SARS-CoV-2 patients (152; 16.2%). Lastly, the Charlson Comorbidity Index showed significant differences (*p* < 0.001), being highest in VRS admissions (3.3 ± 3.1) and lowest in SARS-CoV-2 patients (2.1 ± 2.9).

Symptom presentation varied significantly among different RVIs (*p* ≤ 0.007). Pneumonia was more frequent in VRS (20; 15.2%), and less common in SARS-CoV-2 (81; 8.7%) and influenza (34; 6.8%) patients. LRTI and URTI where significantly more frequently (p= 0.007 and *p*<0.001, respectively) for VRS admissions.

Regarding treatments, significant differences were observed in the use of antibiotics and antivirals among the admissions where the main viruses are presented (*p* < 0.001). Antibiotic use was higher in VRS (83; 62.9%), and lower in SARS-CoV-2 patients (257; 27.5%). Antiviral use was significantly higher in influenza (162; 32.4%), and lower in SARS-CoV-2 (18; 1.9%) and VRS (8; 6.1%) cases.

### Assessing the Effect of Factors on Episode Severity

We assessed the impact of various severity-related factors (ICU, exitus) using a multi-state model. Our dataset defined three states: Admitted, ICU and exitus. All patients started in the Admitted state, with some transitioning to ICU and others directly to exitus. Additionally, transitions from ICU to exitus are possible (**Figure 3**).

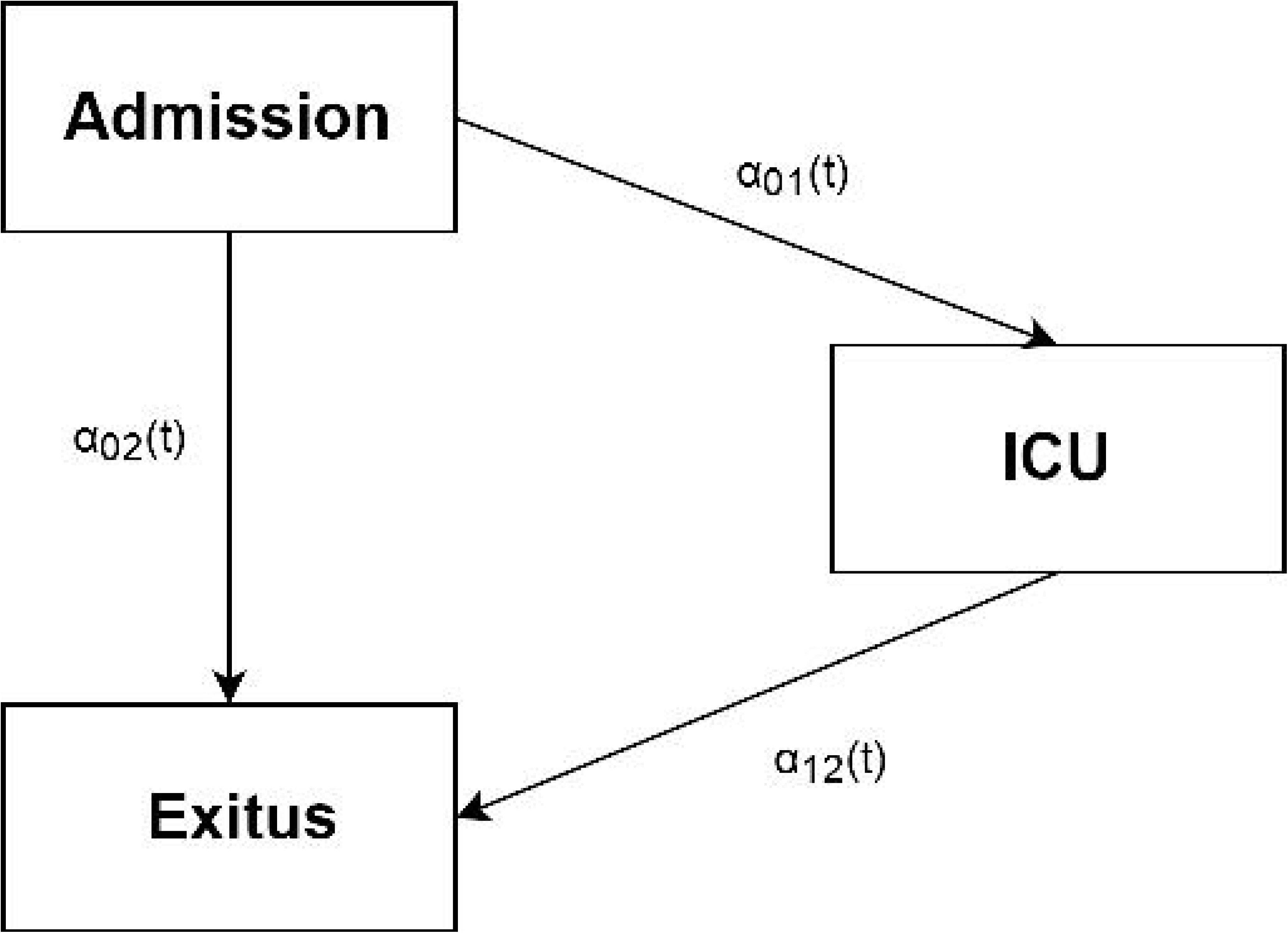

Prior to constructing the multi-state Cox model, we checked whether each univariate predictor variable available was significant (*p*<0.05), as well as assessing whether they met the proportional hazards criterion. In addition, it was assessed whether the continuous variables followed a non-linear distribution or were time dependent. In neither case were these assumptions made (*p*>0.05). After selecting the most relevant variables, we proceeded with the multi-state Cox model analysis (**Table 3**).

**Table 3.** Cox Multistate Inpatient-ICU-exitus Model.

| Item | Admission-ICU Transition |  | Admission -Exitus Transition |  | ICU-Exitus Transition |  |
| --- | --- | --- | --- | --- | --- | --- |
|  | Hazard ratios<br>(95% CI) | *p*- value | Hazard ratios<br>(95% CI) | *p*-value | Hazard ratios<br>(95% CI) | *p*- value |
| Age entry | 0.9 (0.9 - 0.9) | **<0.001** | 1.2 (1.1 - 1.3) | **<0.001** | 1.1 (1.0 - 1.2) | **<0.001** |
| Neoplasia | 0.9 (0.3 - 2.6) | 0.854 | 0.5 (0.2 - 1.4) | 0.203 | 0.5 (0.1 - 3.0) | 0.436 |
| Kidney<br>insufficiency | 0.8 (0.4 - 1.6) | 0.596 | 0.9 (0.4 - 2.1) | 0.860 | 1.9 (0.4 - 8.6) | 0.406 |
| Heart_failure | 1.9 (0.8 - 4.5) | 0.159 | 0.5 (0.21 - 1.4) | 0.200 | 0.8 (0.1 - 4.5) | 0.797 |
| Charlson score | 0.9 (0.8 - 1.1) | 0.274 | 1.4 (1.2 - 1.6) | **<0.001** | 1.6 (0.9 - 2.8) | 0.081 |
| Pneumonia | 0.7 (0.4 - 1.4) | 0.311 | 1.2 (0.5 - 3.2) | 0.689 | 0.4 (0.1 - 2.9) | 0.364 |
| Antibiotics | 0.5 (0.2 - 0.9) | **0.041** | 0.6 (0.0 - 9.0) | 0.710 | 3.7 (0.5 - 25.5) | 0.186 |
| Anticoagulants | 1.9 (0.5 - 7.1) | 0.315 | 0.1 (0.0 - 0.3) | **0.001** | 0.3 (0.0 - 2.7) | 0.308 |
| Diuretics | 0.8 (0.3 - 1.6) | 0.472 | 0.2 (0.1 - 0.9) | **0.038** | 0.2 (0.0 - 2.0) | 0.151 |
| Influenza | 1.1 (0.6 - 2.1) | 0.723 | 1.6 (0.6 - 4.3) | 0.365 | 0.8 (0.2 - 3.8) | 0.783 |
| RSV | 2.2 (1.1 - 4.6) | **0.037** | 0.9 (0.3 - 3.3) | 0.895 | NA (NA - NA) | NA |

CI: Confidence Intervals. Significant differences ( $p < 0.05$ ) are indicated in bold. NA: Not available.

Among all evaluated variables, age at admission showed significant differences across all transitions. For the transition from Admitted to ICU HR was 0.9 (95% CI: 0.9 – 0.9, *p* < 0.001), indicating that older age decreases the likelihood of transfer to ICU. Conversely, for the transition from admission to exitus, HR was 1.2 (95% CI: 1.1 – 1.3, *p* < 0.001), suggesting that older age increases the likelihood of mortality directly from the admission state. Similarly, for the transition from ICU to exitus, HR was 1.1 (95% CI: 1.0 – 1.2, *p* < 0.001), indicating a significantly higher risk of death in ICU with increasing age.

Furthermore, the presence of RSV proved to be a significant risk factor in the transition from admission to ICU with an HR of 2.2 (95% CI: 1.1 – 4.3, *p* = 0.037), indicating that RSV presence increases the likelihood of ICU admission 2.2 times compared to those without RSV. Meanwhile, the presence of antibiotics were a protective factor with an HR of 0.5 (95% CI: 0.2 – 0.9, *p* = 0.041). In the transition from admission to exitus, the presence of a higher score increased a 40% the likelihood to exitus from admission with an HR of 1.4 (95% CI: 1.2 – 1.6, *p* < 0.001). Anticoagulants and diuretic treatment were significant protective factors in the transition from admission to exitus with an HR of 0.1 (95% CI: 0.0 – 0.3, *p* = 0.001) and with an HR of 0.3 (95% CI: 0.1 – 0.9, *p* = 0.038), respectively.

## Discussion

This study offers a comprehensive analysis of RVIs in adults aged 60 and older, emphasising their significant impact on morbidity and mortality in this demographic. Our research identifies SARS-CoV-2, influenza and RSV as the primary viral agents leading to hospitalizations due to respiratory conditions in this cohort. This analysis sheds light on the evolving landscape of viral infections and their implications for public health strategies targeting older adults.

The high incidence of SARS-CoV-2 observed in our sample can be attributed to the analysis period, which followed the COVID-19 epidemics, during which virus testing was paramount. The widespread testing facilitated the early detection and isolation of infected individuals, thereby influencing hospitalization rates. However, this study goes beyond the immediate effects of SARS-CoV-2 to examine the broader spectrum of viral infections affecting older adults.

Our findings reveal a substantial impact of RSV, accounting for 11.8% of respiratory infections in admissions for adults aged 60 and older. This incidence rate of RSV in our study is notably higher than the national average of 7.9% reported in 2024 (33), making it the third most common viral cause after SARS-CoV-2 and influenza. The heightened incidence of RSV underscores the need for increased awareness and targeted interventions to address this often-overlooked virus.

RSV, traditionally considered less impactful in older adults, is associated with higher complication rates during hospitalization, such as pneumonia and LRTIs. These complications frequently require additional treatments, including antibiotics and diuretics. Notably, patients with RSV experienced longer hospital stays and higher mortality rates compared to those with SARS-CoV-2 and influenza. Furthermore, RSV infection significantly increased the risk of ICU admission, with a 2.2-fold higher likelihood of transfer to intensive care.

The impact of RSV is particularly pronounced in older adults with specific risk factors, such as smoking and comorbidities like COPD and heart failure. Advanced age and underlying health conditions significantly elevate the risk of severe disease, highlighting the need for tailored healthcare strategies for these vulnerable groups (5,13).

Respiratory viral infections impose a substantial socioeconomic burden due to increased healthcare utilization and associated costs. While scientific focus has historically been on influenza and SARS-CoV-2, other respiratory viruses like RSV can be equally or more detrimental, particularly in older adults (5,9,13–15). The circulation and aggressiveness of these viruses have shifted post-COVID-19, with notable changes in their seasonality (21,34). This necessitates robust epidemiological research and adaptive immunization strategies to protect the most vulnerable populations effectively.

One of the strengths of this study is its focus on the effects of RSV in adults aged 60 and older in Spain, a relatively underexplored area. This provides significant value as we demonstrate that older adults, particularly those with specific comorbidities, are susceptible and sometimes more vulnerable to RSV than to other more widely recognized viruses. However, the study also has limitations. The use of varied detection techniques for respiratory viruses, such as PCR and antigen tests, might affect the heterogeneity of detection sensitivity and specificity (35). Additionally, these detection tests primarily target the major viruses evaluated in this study and may overlook co-infections with less common viruses. There is a possibility that a higher percentage of co-infections were diagnosed in more severely ill patients, who underwent more extensive testing due to their worse prognosis.

Further research is essential to confirm whether there is a higher morbidity and mortality associated with these viruses. This study reinforces the need to investigate the aetiology of respiratory infections in older adults to identify factors associated with disease severity. Early diagnosis and treatment, coupled with appropriate preventive strategies, could significantly improve healthcare quality, reduce morbidity and mortality, and decrease healthcare resource utilization. Effective public health policies and continuous surveillance are vital for protecting older adults from the evolving threat of respiratory viral infections (12,26). Given the availability of vaccines to prevent severe COVID-19, influenza, and RSV from 2024, public health decisions will be crucial. Future strategies will be instrumental in preventing severe RSV infections in this population. It is essential to emphasize the importance of vaccination, especially since these vaccines are now available in Spain.

## Data Availability

All data produced in this study are available upon reasonable request from the authors

## Institutional Review Board Statement

The study was conducted in accordance with the Declaration of Helsinki, and approved by the Ethics Committee of HM Hospitales (protocol code 24.04.2344-GHM and date of approval: 2024/05/08).” for studies involving humans.

## Informed Consent Statement

Moreover, this research is a retrospective study using anonymized data, which involves no direct patient intervention; consequently, individual consent to participate is not necessary.

## Annex

**Table 1.** Rates of Hospital Admissions with Positive Detection of Respiratory Viruses in Patients Older than 60 Years of Age.

| Episode date | virus | Cases with positive PCR | Total of admissions |  |  | Total of admisiones with respiratory pathology |  |  | Total of admissions with positive virus detection |  |  |
| --- | --- | --- | --- | --- | --- | --- | --- | --- | --- | --- | --- |
|  |  |  | Cases | Rate per 10,000 incomes | Log rate | Cases | Rate per 10,000 incomes | Log rate | Cases | Rate per 10,000 incomes | Log rate |
| 01/10/2023 | SARS-CoV-2 | 273 | 3,622 | 753.7 | 6.6 | 456 | 5,986.8 | 8.7 | 339 | 8,053.1 | 9.0 |
| 01/10/2023 | Influenza virus | 26 | 3,622 | 71.8 | 4.3 | 456 | 570.2 | 6.3 | 339 | 767.0 | 6.6 |
| 01/10/2023 | RSV | 5 | 3,622 | 13.8 | 2.6 | 456 | 109.6 | 4.7 | 339 | 147.5 | 5.0 |
| 01/10/2023 | Rhinovirus | 26 | 3,622 | 71.8 | 4.3 | 456 | 570.2 | 6.3 | 339 | 767.0 | 6.6 |
| 01/10/2023 | Other respiratory viruses | 9 | 3,622 | 24.8 | 3.2 | 456 | 197.4 | 5.3 | 339 | 265.5 | 5.6 |
| 01/10/2023 | Metapneumovirus | 0 | 3,622 | 0.0 | 0.0 | 456 | 0.0 | 0.0 | 339 | 0.0 | 0.0 |
| 01/11/2023 | SARS-CoV-2 | 141 | 3,841 | 367.1 | 5.9 | 430 | 3,279.1 | 8.1 | 316 | 4462 | 8.4 |
| 01/11/2023 | Influenza virus | 52 | 3,841 | 135.4 | 4.9 | 430 | 1,209.3 | 7.1 | 316 | 1,645.6 | 7.4 |
| 01/11/2023 | RSV | 46 | 3,841 | 119.8 | 4.8 | 430 | 1,069.8 | 7.0 | 316 | 1,455.7 | 7.3 |
| 01/11/2023 | Rhinovirus | 45 | 3,841 | 117.2 | 4.8 | 430 | 1,046.5 | 7.0 | 316 | 1,424.1 | 7.3 |
| 01/11/2023 | Other respiratory viruses | 30 | 3,841 | 78.1 | 4.4 | 430 | 697.7 | 6.5 | 316 | 949.4 | 6.9 |
| 01/11/2023 | Metapneumovirus | 2 | 3,841 | 5.2 | 1.6 | 430 | 46.5 | 3.8 | 316 | 63.3 | 4.1 |
| 01/12/2023 | SARS-CoV-2 | 307 | 3,224 | 952.2 | 6.9 | 908 | 3,381.1 | 8.1 | 795 | 3,861.6 | 8.3 |
| 01/12/2023 | Influenza virus | 282 | 3,224 | 874.7 | 6.8 | 908 | 3,105.7 | 8.0 | 795 | 3,547.2 | 8.2 |
| 01/12/2023 | RSV | 77 | 3,224 | 238.8 | 5.5 | 908 | 848 | 6.7 | 795 | 968.6 | 6.9 |
| 01/12/2023 | Rhinovirus | 64 | 3,224 | 198.5 | 5.3 | 908 | 704.8 | 6.6 | 795 | 805 | 6.7 |
| 01/12/2023 | Other respiratory viruses | 57 | 3,224 | 176.8 | 5.2 | 908 | 627.8 | 6.4 | 795 | 717 | 6.6 |
| 01/12/2023 | Metapneumovirus | 8 | 3,224 | 24.8 | 3.2 | 908 | 88.1 | 4.5 | 795 | 100.6 | 4.6 |
| 01/01/2024 | SARS-CoV-2 | 208 | 4,037 | 515.2 | 6.2 | 818 | 2,542.8 | 7.8 | 591 | 3,519.5 | 8.2 |
| 01/01/2024 | Influenza virus | 220 | 4,037 | 545.0 | 6.3 | 818 | 2,689.5 | 7.9 | 591 | 3,722.5 | 8.2 |
| 01/01/2024 | RSV | 70 | 4,037 | 173.4 | 5.2 | 818 | 855.7 | 6.8 | 591 | 1,184.4 | 7.1 |
| 01/01/2024 | Rhinovirus | 27 | 4,037 | 66.9 | 4.2 | 818 | 330.1 | 5.8 | 591 | 456.9 | 6.1 |
| 01/01/2024 | Other respiratory viruses | 53 | 4,037 | 131.3 | 4.9 | 818 | 647.9 | 6.5 | 591 | 896.8 | 6.8 |
| 01/01/2024 | Metapneumovirus | 13 | 4,037 | 32.2 | 3.5 | 818 | 158.9 | 5.1 | 591 | 220.0 | 5.4 |
| 01/02/2024 | SARS-CoV-2 | 61 | 3,833 | 159.1 | 5.1 | 300 | 2,033.3 | 7.6 | 129 | 4,728.7 | 8.5 |
| 01/02/2024 | Influenza virus | 23 | 3,833 | 60.0 | 4.1 | 300 | 766.7 | 6.6 | 129 | 1,782.9 | 7.5 |
| 01/02/2024 | RSV | 10 | 3,833 | 26.1 | 3.3 | 300 | 333.3 | 5.8 | 129 | 775.2 | 6.7 |
| 01/02/2024 | Rhinovirus | 15 | 3,833 | 39.1 | 3.7 | 300 | 500.0 | 6.2 | 129 | 1,162.8 | 7.1 |
| 01/02/2024 | Other respiratory viruses | 3 | 3,833 | 7.8 | 2.1 | 300 | 100.0 | 4.6 | 129 | 232.6 | 5.4 |
| 01/02/2024 | Metapneumovirus | 17 | 3,833 | 44.4 | 3.8 | 300 | 566.7 | 6.3 | 129 | 1,317.8 | 7.2 |
| 01/03/2024 | SARS-CoV-2 | 26 | 3,644 | 71.4 | 4.3 | 346 | 751.4 | 6.6 | 115 | 2,260.9 | 7.7 |
| 01/03/2024 | Influenza virus | 29 | 3,644 | 79.6 | 4.4 | 346 | 838.2 | 6.7 | 115 | 2,521.7 | 7.8 |
| 01/03/2024 | RSV | 21 | 3,644 | 57.6 | 4.1 | 346 | 606.9 | 6.4 | 115 | 1,826.1 | 7.5 |
| 01/03/2024 | Rhinovirus | 17 | 3,644 | 46.7 | 3.8 | 346 | 491.3 | 6.2 | 115 | 1,478.3 | 7.3 |
| 01/03/2024 | Other respiratory viruses | 8 | 3,644 | 22.0 | 3.1 | 346 | 231.2 | 5.4 | 115 | 695.7 | 6.5 |
| 01/03/2024 | Metapneumovirus | 14 | 3,644 | 38.4 | 3.6 | 346 | 404.6 | 6.0 | 115 | 1,217.4 | 7.1 |

**Table 2.** Admission (A), ICU admission (B) and mortality (C) rates and 95% confidence intervals of patients aged 60 years and older for RVIs during the from 1 October 2023 to 31 March 2024. Rates are expressed per 10,000 admission, ICU or Deceased admissions patients with these characteristics, and

| Group | Age range | SARS-CoV-2<br>Estimator [CI<sub>95%</sub>] | Mean<br>rate | Influenza virus<br>Estimator [CI<sub>95%</sub>] | Mean<br>rate | Respiratory syncytial<br>virus<br>Estimator [CI<sub>95%</sub>] | Mean<br>rate | Rhinovirus<br>Estimator [CI<sub>95%</sub>] | Mean<br>rate |
| --- | --- | --- | --- | --- | --- | --- | --- | --- | --- |
| Total admissions | 60-70 years | 333.4 [295.0 - 375.2]* | 518.9 | 169.8 [142.6 - 200.7]* | 347.1 | 69.2 [52.2 - 90.0]* | 139.7 | 61.6 [45.6 - 81.4]* | 107.4 |
| Total admissions | 70-79 years | 425.5 [382.6 - 471.6]* | 518.9 | 273.0 [238.7 - 310.8]* | 347.1 | 70.1 [53.1 - 90.7]* | 139.7 | 76.2 [58.5 - 97.6] | 107.4 |
| Total admissions | 80-89 years | 665.1 [594.9 - 740.8]* | 518.9 | 426.8 [370.3 - 489.1]* | 347.1 | 173.3 [137.7 - 215.2] | 139.7 | 125.7 [95.6 - 162.1] | 107.4 |
| Total admissions | ≥90 years | 651.6 [532.1 - 788.4]* | 518.9 | 518.6 [412.1 - 643.1]* | 347.1 | 246.0 [173.8 - 337.5]* | 139.7 | 166.2 [107.9 - 244.4] | 107.4 |
| ICU admissions | 60-70 years | 84.8 [36.7 - 166.3] | 86.7 | 84.8 [36.7 - 166.3] | 127.1 | 53.0 [17.2 - 123.2] | 41.3 | 53.0 [17.2 - 123.2] | 35.6 |
| ICU admissions | 70-79 years | 101.4 [52.5 - 176.4] | 86.7 | 118.2 [64.8 - 197.6] | 127.1 | 33.8 [9.2 - 86.3] | 41.3 | 16.9 [2.1 - 60.9] | 35.6 |
| ICU admissions | 80-89 years | 74.1 [20.2 - 188.6] | 86.7 | 166.7 [76.5 - 314.0] | 127.1 | 37.0 [4.5 - 133.1] | 41.3 | 37.0 [4.5 - 133.1] | 35.6 |
| ICU admissions | ≥90 years | -- | -- | 138.9 [3.5 - 749.7] | 127.1 | -- | -- | -- | -- |
| Deceased admissions | 60-70 years | 333.3 [40.6 - 1152.8] | 320.3 | 333.3 [40.6 - 1152.8] | 333.5 | -- | -- | 333.3 [40.6 - 1152.8] | 185.5 |
| Deceased admissions | 70-79 years | 279.7 [76.7 - 700.7] | 320.3 | 349.7 [114.5 - 797.1] | 333.5 | 69.9 [1.8 - 383.5] | 173.5 | 139.9 [17.0 - 496.1] | 185.5 |
| Deceased admissions | 80-89 years | 304.4 [123.2 - 617.0] | 320.3 | 347.8 [151.3 - 673.8] | 333.5 | 87.0 [10.6 - 310.6] | 173.5 | 87.0 [10.6 - 310.6] | 185.5 |
| Deceased admissions | ≥90 years | 363.6 [134.6 - 774.7] | 320.3 | 303.0 [99.1 - 693.0] | 333.5 | 363.6 [134.6 - 774.7] | 173.5 | 181.8 [37.7 - 522.1] | 185.5 |

CI: Confidence interval. The 95% CI that did not include the mean rate value were marked with an asterisk and in bold.

